# Trust in data sharing reflects the contextuality of the trustor–trustee relationship

**DOI:** 10.1101/2025.07.05.25330938

**Authors:** Jonathan R Goodman, Alessia Costa, Richard Milne

**Affiliations:** Wellcome Sanger Institute, Cambridge, UK; Cambridge Public Health, University of Cambridge, UK; Kavli Centre for Ethics Science and the Public, University of Cambridge, UK

## Abstract

It is widely recognized that trust is an essential element in how people engage with data sharing and underpins efforts to use data to improve care quality and understand population-level health trends, and consequently improve health inequalities. However, research into public trust in the data sharing and healthcare settings may rely on oversimplified notions of what trust entails, and what is involved in trusting relationships in this setting has not been widely explored. Relatedly, the way that trust manifests as a function of perceived trustworthiness is unestablished. Here, we analyze data from 2000 participants who completed a questionnaire about how they place trust in entities including family, the healthcare system, and corporations when it comes to their personal health data. We find that the reasons people place trust differ depending on the relationship and circumstance, and suggest that trustworthiness is an underlying quality that manifests and is perceived differently under different conditions. Future work into trust relationships should account for this varying presentation of trust, and perceptions of trustworthiness, when it comes to exploration of relationships between people, institutions, and systems.

## Introduction

Trust in the institutions involved in health research and care is consequential, shaping decisions around the willingness to make data available for secondary uses including research (Middleton et al., 2019). Given the growing interest in the use of digital and data-driven tools in medicine and public health, this has potential wider implications for the ability to deliver effective, equitable services.

In the literature exploring individual-level decisions to share data related to health, researchers tend to place emphasis on the entity being trusted. For example, numerous studies have focused on whether individuals are more likely to share their data with healthcare practitioners, government bodies, corporations, universities, and so forth (e.g. Baines et al., 2024; Kalkman et al., 2022). Work on trust in scientific and healthcare institutions is, consequently, broadly concerned with why individuals place trust in specific entities and not others — with emphasis placed on population-level differences in trust placement, such as between countries or between groups within populations — including, for example, between groups based on ethnicity and socioeconomic status (Middleton et al., 2023; Milne et al., 2019a).

To date, however, the literature on trust in health data has engaged in less detail with what is involved in public trust in data institutions (including the NHS and health technology companies), what members of the public associate with the trustworthiness of these institutions (Ghafur et al., 2020; O. Lounsbury et al., 2021), and how this relates to the current or future adoption of digital health technologies. In this paper, focus on the qualities or characteristics that are associated with the perceived trustworthiness of the institutions involved in collecting and working with data about health, and the relationship of these qualities with those of trustors.

### Institutions that are worthy of trust

A focus on public trust in institutions that collect, use, and share health data implicitly treats these institutions implicitly or explicitly as having qualities that are associated (or not) with being worthy of trust. Accordingly, researchers interested in trust, have aimed to determine what makes an institution trustworthy — assuming trustworthiness at an institutional level makes sense (Lounsbury, 2023; Milne et al., 2021). Qualities associated with trustworthiness in this literature include considerations such as familiarity, for both trustor and trustee (Hardin, 2002; Luhmann, 1979), honesty and openness (O’Neill, 2018), competence (D’Cruz, 2019) and reliability (Hawley, 2019), shared values or encapsulated interests (Hardin, 2006), good intentions or goodwill (Baier, 1986), and responsibility (Holton, 1994 — “do they act as they should?”).

The aim in this context is not to go into detail on the merits or otherwise of these accounts of trustworthiness, but to describe how they highlight a range of qualities, behaviours and dispositions that are potentially related to trustworthiness, though with little empirical work to unpick their role in everyday life or in how they enable trustworthy institutions and people to distinguish themselves from bad actors (Goodman & Milne, 2024). While this movement from philosophical conceptualisation into empirical survey presents challenges in terms of the loss of nuance and the openness to interpretation, this literature does offer carefully structured categories that can provide a scaffold for empirical investigation while connecting empirical questions about trust in practice to the normative accounts proposed by Hawley, O’Neill, and colleagues (cf Hawley, 2014) Following Dewey’s pragmatist and pluralistic approach (Sorrell, 2013), the normative concepts of trust can be taken as working hypotheses to be tested, operationalised and refined in real-world contexts through social research to examine how members of the public interpret and prioritize different grounds for trust.

Indeed, from the existing literature, it is an open question whether or how philosophical accounts of trust accord with how people place trust in the real world. For example, transparency around data usage does not appear, in practice, to have the implicit moral content assumed by researchers working in the social sciences and humanities (Laverty et al., 2024). Instead it may be linked with negative sentiment insofar as members of the public are not empowered to act on the information they are given about how their data are used. This accords with work by Jones (2012) suggesting that desirable qualities, including those that constitute trustworthiness, are not necessarily useful if they are not observable: it is no benefit to be trustworthy if no one can distinguish you from bad actors (see Goodman and Milne, 2024 for a discussion of the trustworthiness recognition problem). Being able to recognise the trustworthy is essential to be able to place trust ‘intelligently’ (O’Neill 2018).

Moreover, it is not known whether the qualities people associate with trustworthiness vary with the type of entity seeking trust. While transparency may be a requirement for trust among one entity, such as a government, it does not follow that it is required for trust in other circumstances, such as within a family (John, 2021). This potential inconsistency among desired qualities in trustees suggests that normative discourse around trustworthiness, visibility, transparency, and so forth are unlikely to capture important elements of the contextual relationship between trustor and trustee. The manner in which the relationship between trustor and trustee manifests is also likely to drive behaviours at the individual level. These behaviours may not, furthermore, match academic intuitions about how people categorized into low-or high-trust groups are likely to act, just as increased transparency may not lead to increased trust (Felzmann et al., 2019).

Instead, investigating how members of the public self-describe the way they trust, and under what circumstances, and the link of trust relationships between self-reported behaviours, is necessary for revealing whether current uses of terms like “trustworthiness” and “transparency” in the literature predict real-world behavioural patterns. In line with these insights, we conducted a public survey to explore the challenges associated with trust and judgements of trustworthiness in the context of the use of technology to collect health-related data. This context reflects the growing interest in the use of digital devices to collect health information, and the challenges this presents to the distribution of health data between the public and private sectors. Specifically, we aimed to answer the following questions:

1. Which are the primary reasons for the placement of trust, and do these reasons vary by demographics and domain?
2. Does stated general trust, and do demographic characteristics, predict for a composite domain-specific trust score?
3. Does a composite trust score predict for use of technology in the health sphere?
4. Is trust in tech companies a predictor for using technology for health reasons?
5. Is trust in England’s National Health Service (NHS) a predictor for using technology for health reasons?

We hypothesized, in line with previous work, that trust is likely to vary by domain (Stewart, 2024), but also by demographic qualities of the trustor — and that the way trust is placed is likely to predict whether and how people use data-driven health technologies. If correct, this hypothesis may indicate that trustworthiness manifests and is perceived differently under different conditions, and consequently should be understood as a function of the context in which the term is used.

## Methods

### Survey design and data collection

We designed the survey on the Qualtrics (https://www.qualtrics.com) platform and used the marketing firm, Dynata (https://www.dynata.com), to recruit a sample representative of adults living in the United Kingdom on education, sex, and age. The full questionnaire is available in the online supplementary material, and was designed to explore both the likelihood of a person placing trust generally (using a five point version of the general trust question; (OECD Guidelines on Measuring Trust, 2017), and in a given entity in relation to data about their health (family, the NHS, health technology companies). The latter two of these were chosen to reflect the organisations involved in collecting and using health data, while the former represents those proximal to the data subject. Participants were asked to rate their level of trust in others generally (“Generally speaking, would you say that people can be trusted?”) and in specific entities (“Do you feel that you can generally trust the NHS with data about your health?”) on an ordinal scale (1-5), which included “Definitely not,” “Probably not,” “Might or might not,” “Probably,” and “Definitely,” respectively.

We also explored the reasons that people place trust generally or specifically in relation to health data in their family, the NHS, or in health technology companies, which could be any or all of the following:

– “They do what they do with the best of intentions.”
– “They are good at what they do.”
– “They behave responsibly.”
– “They are reliable and keep their promises.”
– “They would not try to take advantage of me if they could.”
– “They are open about what they are doing.”
– “They share my values.”
– “They are familiar to me.”

These questions draw on previously published research aiming to explain the placement of trust (Ipsos Global Advisor, 2018), informed and adapted to reflect the diverse conceptual literature around trustworthiness. In the widely adopted ABI model of trust (Mayer et al., 1995), for example, trustworthiness has often been associated with ability (good at what they do), benevolence (best of intentions), and integrity (behave responsibly, would not take advantage). As discussed above, the wider sociological and philosophical literature highlights considerations such as familiarity, honesty and openness, competence and reliability, shared values or encapsulated interests, good intentions or goodwill, and responsibility.

We also asked whether participants were likely to either use, or whether they owned, technological tools that could theoretically be used to analyse personal health data or provide tailored information about one’s health. Finally, we collected demographic data, which included age, gender, education, caring status, relationship status, and whether the participant had children. The study received research ethics approval at the Faculty of Education at the University of Cambridge in July 2022. We received written consent for all participants, which was obtained online by Dynata; recruitment was conducted between July 20-22^nd^, 2022.

### Analysis

We evaluated the demographic data and general responses to the online questionnaire using base functions in R using R Studio (R Core Team, 2022) and created visualizations of the data using the ggplot2 package for R (Wickham et al., 2021). We then created linear models in a Bayesian framework using the brms package (Bürkner et al., 2022), relying on Markov Chain Monte Carlo (MCMC) sampling. In our first set of models, we evaluated whether participant demographic characteristics were predictive of the likelihood in higher general trust, or in placing trust in family, the NHS, or health technology companies. We visualized the predictions of these models using the sjPlot package for R (Lüdecke et al., 2024).

Next, we evaluated whether the reasons for the placement of trust differed by domain and for general trust. To do so this we plotted the most given reasons for why the participant would place trust in family, the NHS, and health technology companies, and in general. We constructed Bayesian hierarchical models to evaluate whether demographic characteristics predicted for reasons for domain-specific or general trust.

Finally, we created a composite trust score across health data domains (i.e. excluding general trust), where participants could score between 1 and 15. A score of 1 indicates low trust across the domains of family, NHS, and health technology, while a score of 15 indicates very high trust across these domains. We developed our final set of models to determine whether a composite trust score predicts the likelihood of having used, or using, a device to track health or well-being.

All anonymized data and code are available in the study’s online supplementary material.

## Results

### Demographics

Overall, 1192 participants took part in the survey; full demographic data from our sample are available in the online supplementary material and **Table 1**. All participants were aged at least 18 years, with both age ranges and sexes balanced in the sample. Nearly all (99.5%) participants had completed at least secondary schooling, while 60% were married, in a civil partnership, or living with a partner. A small subset of patients (14.6%) self-described as carers.

**Table 1:** Demographic data for the population sample.

|  |  |
| --- | --- |
| **Age** |  |
| 18-24 years old | 145 |
| 25-34 years old | 233 |
| 35-44 years old | 239 |
| 45-54 years old | 225 |
| 55-64 years old | 196 |
| 65+ years old | 153 |
| Unknown | 1 |
| **Sex** |  |
| Male | 587 |
| Female | 605 |
| **Education** |  |
| Primary school | 6 |
| Secondary school | 554 |
| University degree or equivalent | 487 |
| Postgraduate degree | 145 |
| **Relationship status** |  |
| Married/civil partnership/living together | 720 |
| Widowed | 32 |
| Divorced | 75 |
| Separated | 17 |
| Single | 348 |
| **Children** |  |
| Yes | 672 |
| No | 517 |
| **Carer** |  |
| Yes | 174 |
| No | 1015 |

#### 1. Which are the primary reasons for the placement of trust, and do these reasons vary by demographics and domain?

##### General trust

When asked generally whether people can be trusted (“Generally speaking, would you say that people can be trusted?”), the responses were bimodally distributed, with the most frequently chosen answers being “Usually not” and “Usually” (each > 400 out of 1191 total responses; **Figure 1**), according with previous research (Our World in Data, 2022).

**Figure 1:**
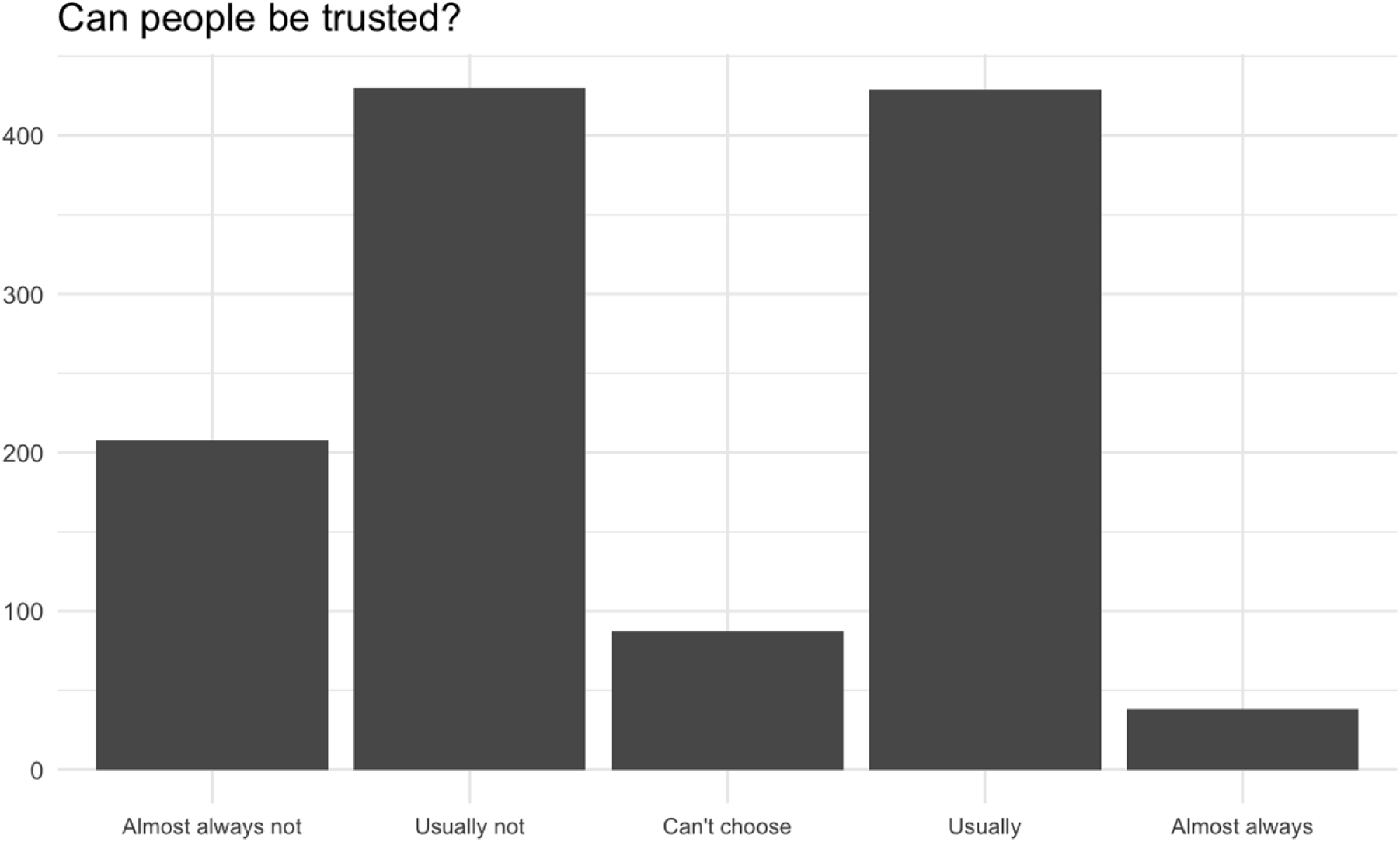
Participants were most likely to indicate people are “usually not” and “usually” worthy of trust, with “almost always not” and “usually not” together composing more than half the total number of responses.

Initial analysis using a Bayesian cumulative logit model suggested that age, gender, and education did not predict for general trust (see the online supplementary material); divorced status, however, predicted for choosing “almost always not” or “usually not” (Beta estimate: –0.71; 95% CI: –1.17 – –0.28; **Figure 2**).

**Figure 2:**
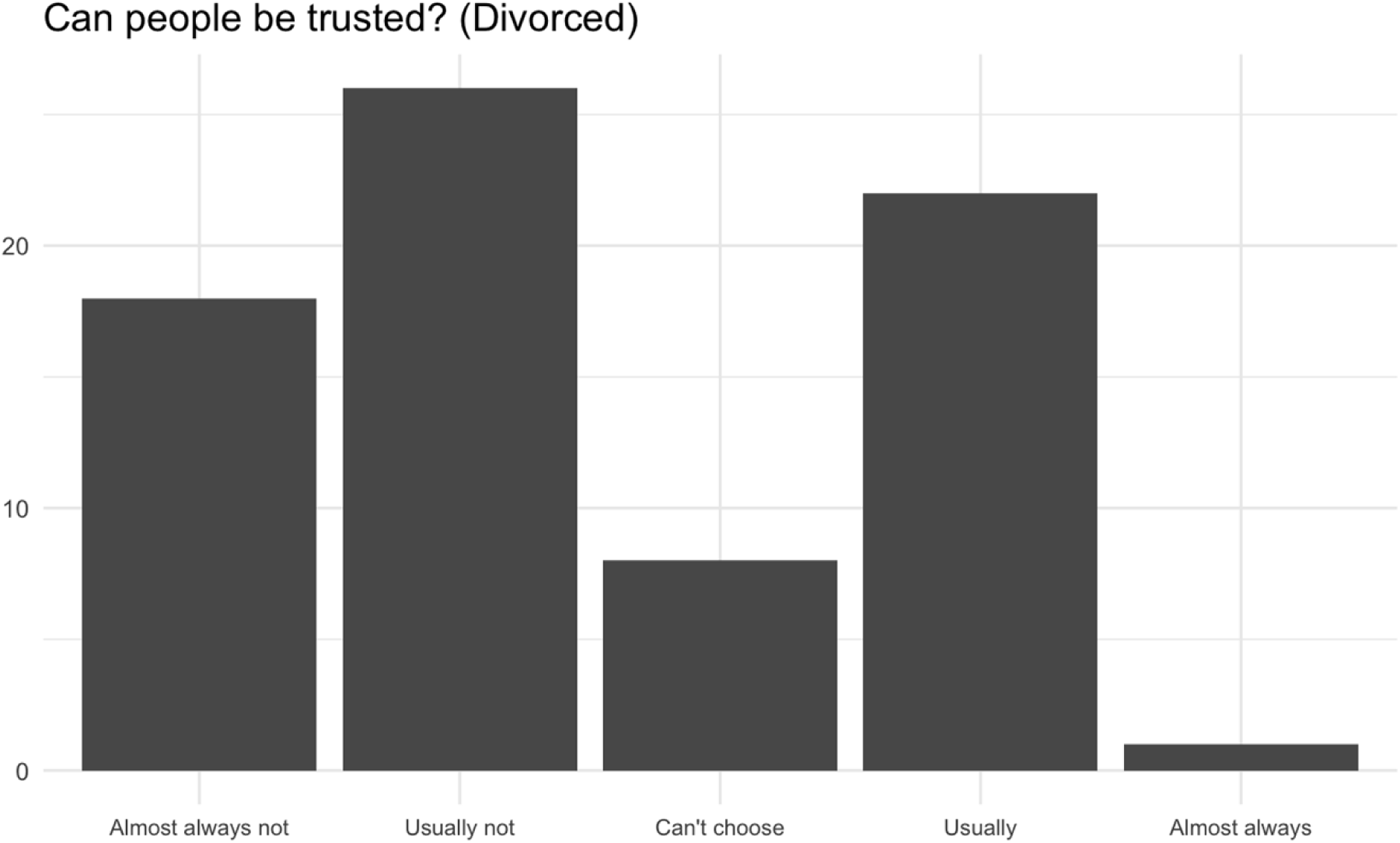
Participants who indicated they were divorced were more likely to indicate people can “almost always not” or “usually not” be trusted (Beta estimate: –0.71; 95% CI: –1.17 – –0.28).

When asked to indicate the three most important reasons for placing trust in others (“In general, when you think about whether you can trust someone, which of these reasons would you say were the three most important?”), the most chosen response was “They are reliable and keep their promises” (771 cases) followed by “They behave responsibly” (629 cases); “They share my values” was chosen least (264 cases; **Figure 3**).

**Figure 3:**
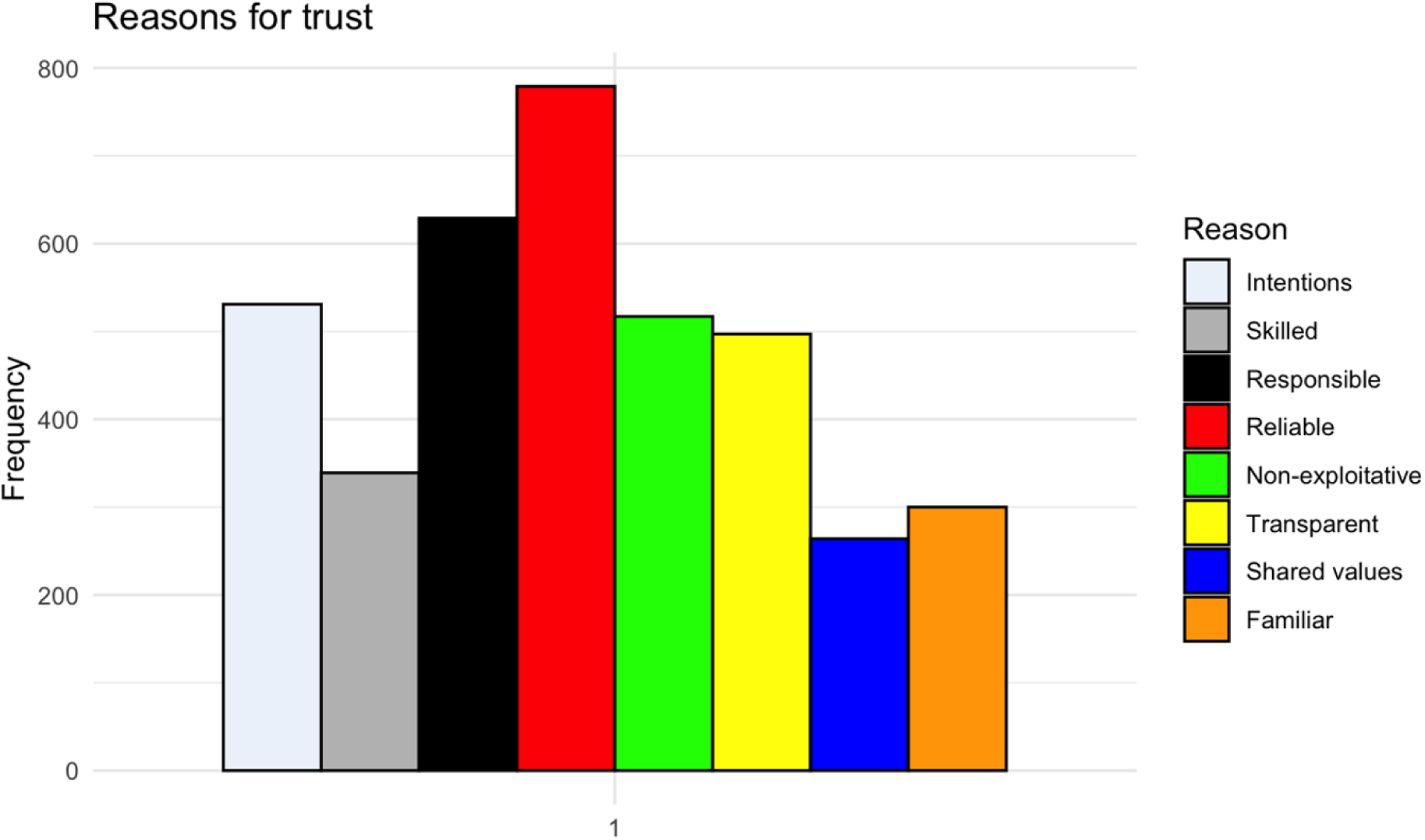
Responses to the question “In general, when you think about whether you can trust someone, which of these reasons would you say were the three most important?” The most chosen response was “They are reliable and keep their promises” (771 cases) followed by “They behave responsibly” (629 cases); “They share my values” was chosen least (264 cases).

A Bayesian hierarchical model using participant as a random effect suggested that most demographic characteristics are not predictive of reasons for the placement of trust, though older age groups were linked with a greater likelihood of choosing “They are familiar to me,” which otherwise received fewer responses from the cohort (see the online supplementary material for details).

##### Trust in family

Most participants indicates that they would “usually” or “almost always” trust family with their health data (**Table 2**). Fewer than 50 participants each, by contrast, chose “almost always not” or “usually not.” A Bayesian cumulative logit model suggested that only single status predicted for a lower trust rating towards family (Beta estimate: –0.39; 95% CI: –0.66 –-0.13; see the online supplementary material).

**Table 2:** stated trust levels in the overall cohort, including general trust, trust in family, trust in the NHS, and trust in health technology companies; all data presented as a percentage of responses.

| Response | General | Family | NHS | Tech companies |
| --- | --- | --- | --- | --- |
| You almost always can’t be too careful in dealing with people | 17.45 | 1.43 | 2.6 | 8.15 |
| You usually can’t be too careful in dealing with people | 36.07 | 3.87 | 6.47 | 15.63 |
| Can’t choose | 7.3 | 14.9 | 14.44 | 31.51 |
| People can usually be trusted | 35.99 | 36.87 | 47.86 | 33.36 |
| People can almost always be trusted | 3.19 | 42.93 | 28.63 | 11.34 |

The reasons primarily chosen for placing trust in family regarding health data (“Thinking about whether your family can be trusted with data about your health, which three of these reasons for trusting would be most important to you?”), participants most frequently chose familiarity (“They are familiar to me”), and least frequently chose “They are good at what they do” (labelled as “Skilled” in **Figure 4**). A Bayesian hierarchical model suggested that older participants (55-64 and 65 years plus) were least likely to select “They are good at what they do,” though the estimates were not strong (see the online supplementary material).

**Figure 4:**
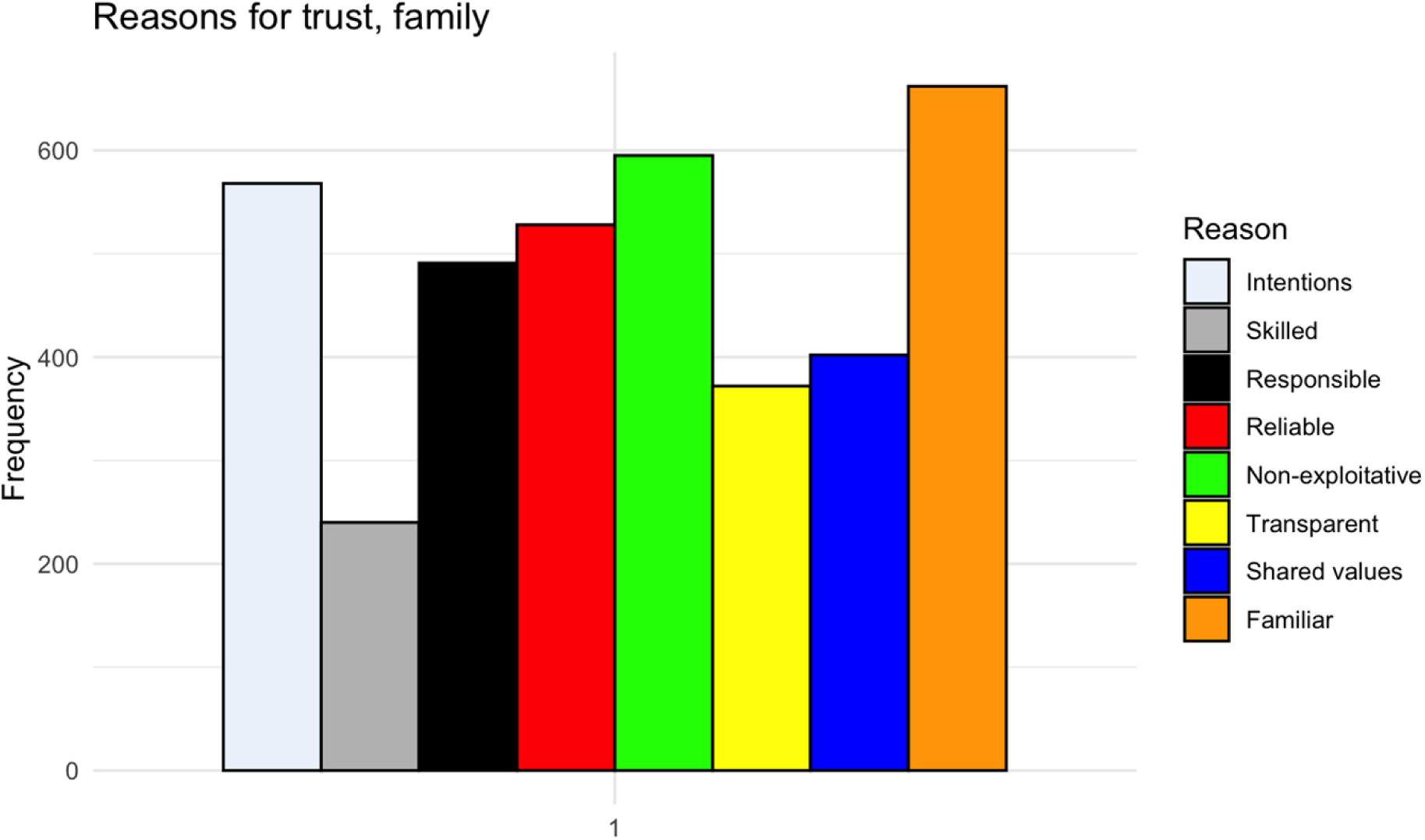
Primary reasons for the placement of trust in family members with health data. “They are familiar to me” and “They would not take advantage of me” were the most frequently chosen responses, while “They are good at what they do” was least frequently chosen.

##### Trust in the NHS

As with family, the majority of participants selected “usually” or “almost always” when asked “Can the NHS be trusted with your health data?” (**Table 2**). Compared with a primary school education, secondary school (Beta estimate: 1.96; 95% CI: 0.64–3.29), university degree or equivalent (Beta estimate: 1.83; 95% CI: 0.51–3.15), and postgraduate degree (Beta estimate: 2.08; 95% CI: 0.71–3.43) predicted for increased trust in the NHS, with similar relative estimates between those groups. “They behave responsibly” was the primary reason chosen for trust in the NHS, while “They share my values” was chosen the least frequently (**Figure 5**); older age groups were less likely to choose “They share my values” (see the online supplementary material).

**Figure 5:**
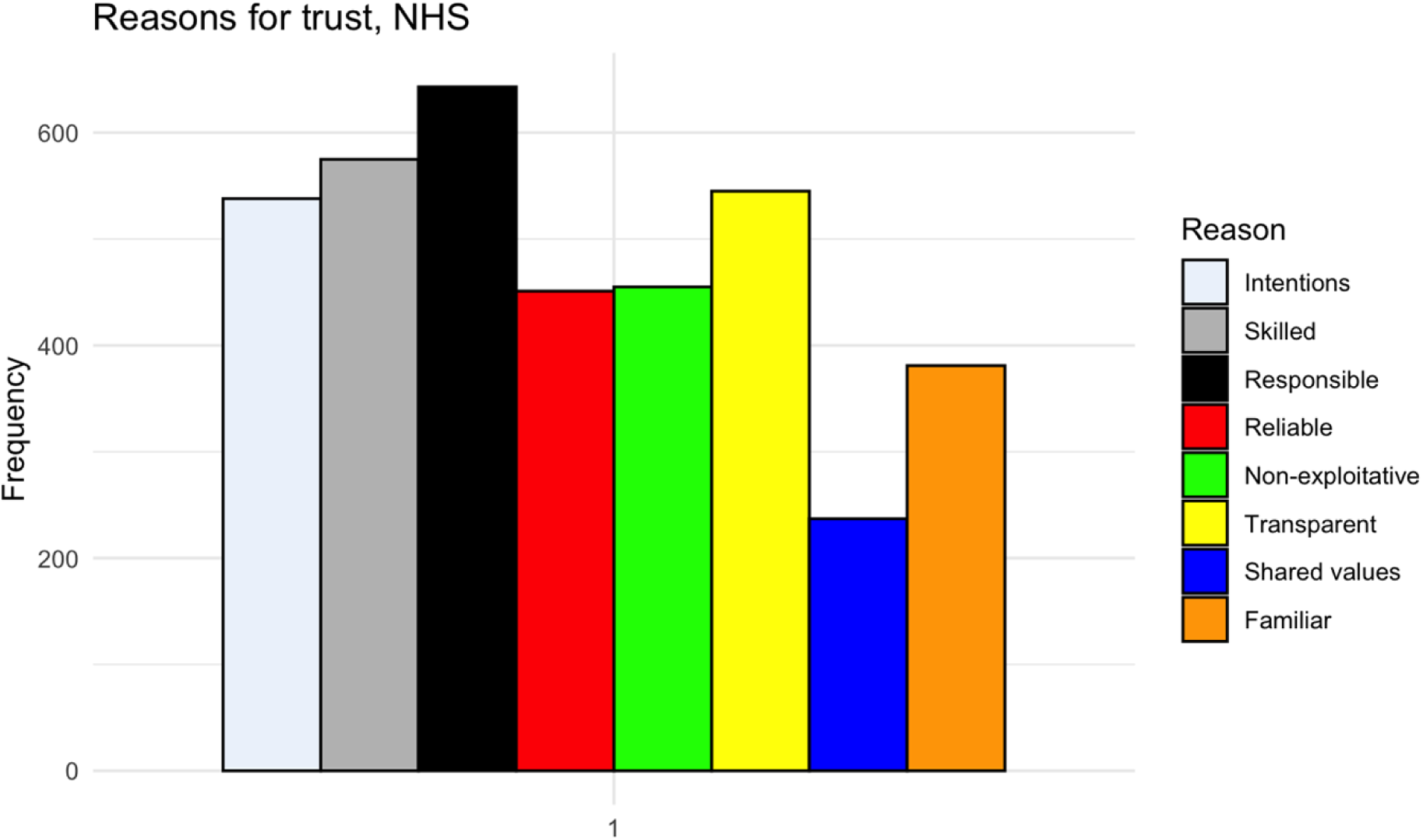
“They behave responsibly” was the most frequently chosen reason for trust in the NHS.

##### Trust in health technology companies

Compared with family and the NHS, fewer participants chose “almost always” when asked “can tech companies be trusted with your health data?” (**Table 2**); most participants chose “usually” followed by “can’t choose.” Single relationship status was a weak predictor for lower trust in these entities (Beta estimate: –0.35; 95% CI: –0.61 – –0.10). “They behave responsibly” and “They are open about what they do” were the most frequently chosen reasons for trust in this setting; “They share my values” was least frequently chosen. A Bayesian hierarchical model evaluating reasons did not suggest any strong relationships between demographic variables and the reasons for trust in health technology companies (**Figure 6**; see the online supplementary material).

**Figure 6:**
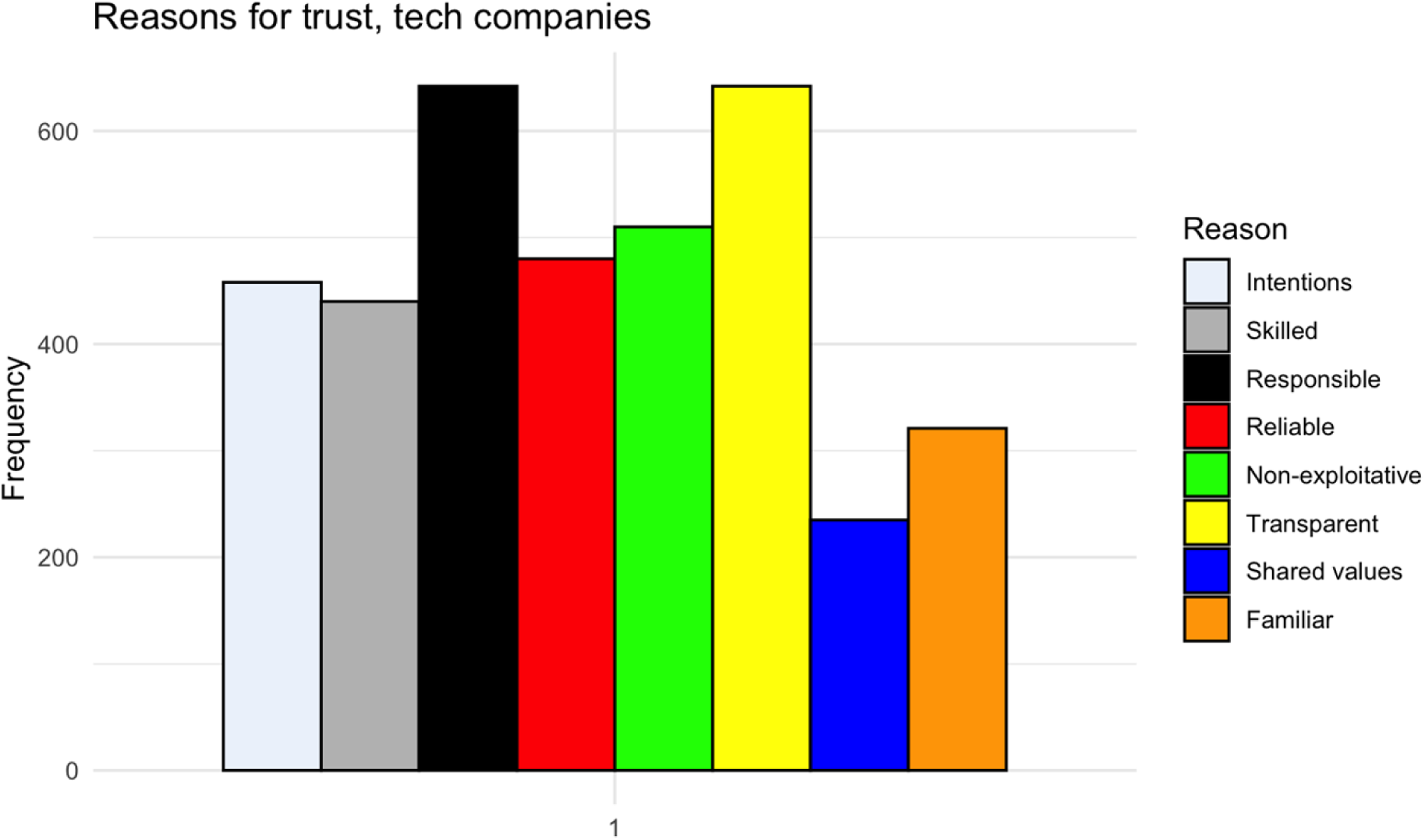
“They behave responsibly” and “They are open about what they do” were the most frequently chosen reasons for trust in health technology companies; “They share my values” was least frequently chosen.

#### 2. Does stated general trust, and do demographic characteristics, predict for a composite domain-specific trust score?

We next created a composite score (1-15; minimum trust = 1; maximum trust = 15) adding together self-scores on the domain-specific questions around trust (family, NHS, health tech companies; each 1-5). A Bayesian cumulative logit model suggested that higher general trust was a strong predictor of a higher composite trust score (Beta estimate for a self-score of 5 on general trust vs 1: 1.93; 95% CI: 1.26–2.59). Single relationship status (Beta estimate: –0.31; 95% CI: –0.56 – –0.07) was the only demographic variable that predicted for the composite trust score, and was both weak and negative.

Categorizing participants by composite score as low trustors (score 1-5; 65 participants), medium trustors (score 6-10; 755 participants), or high trustors (score 11-15; 365 participants) suggested that, across domains, reliability was a highly rated reason for the placement of trust, though low trustors placed slightly more emphasis on responsibility (**Figure 7**). Shared values and familiarity, by contrast, were consistently chosen as the least important reasons across the three groups.

3. *Does a composite trust score predict for use of technology in the health sphere?*
4. *Is trust in tech companies a predictor for using technology for health reasons?*
5. *Is trust in England’s National Health Service (NHS) a predictor for using technology for health reasons?*

**Figure 7:**
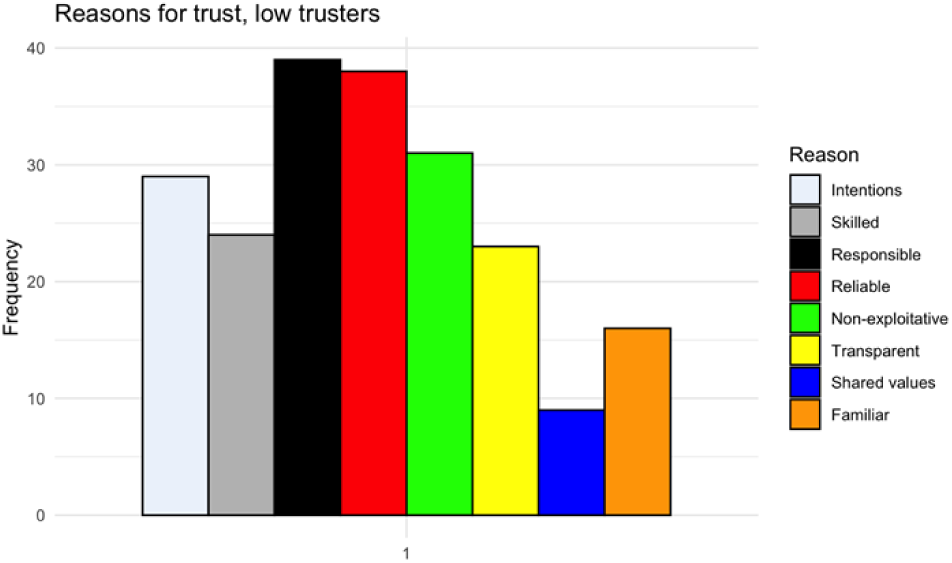
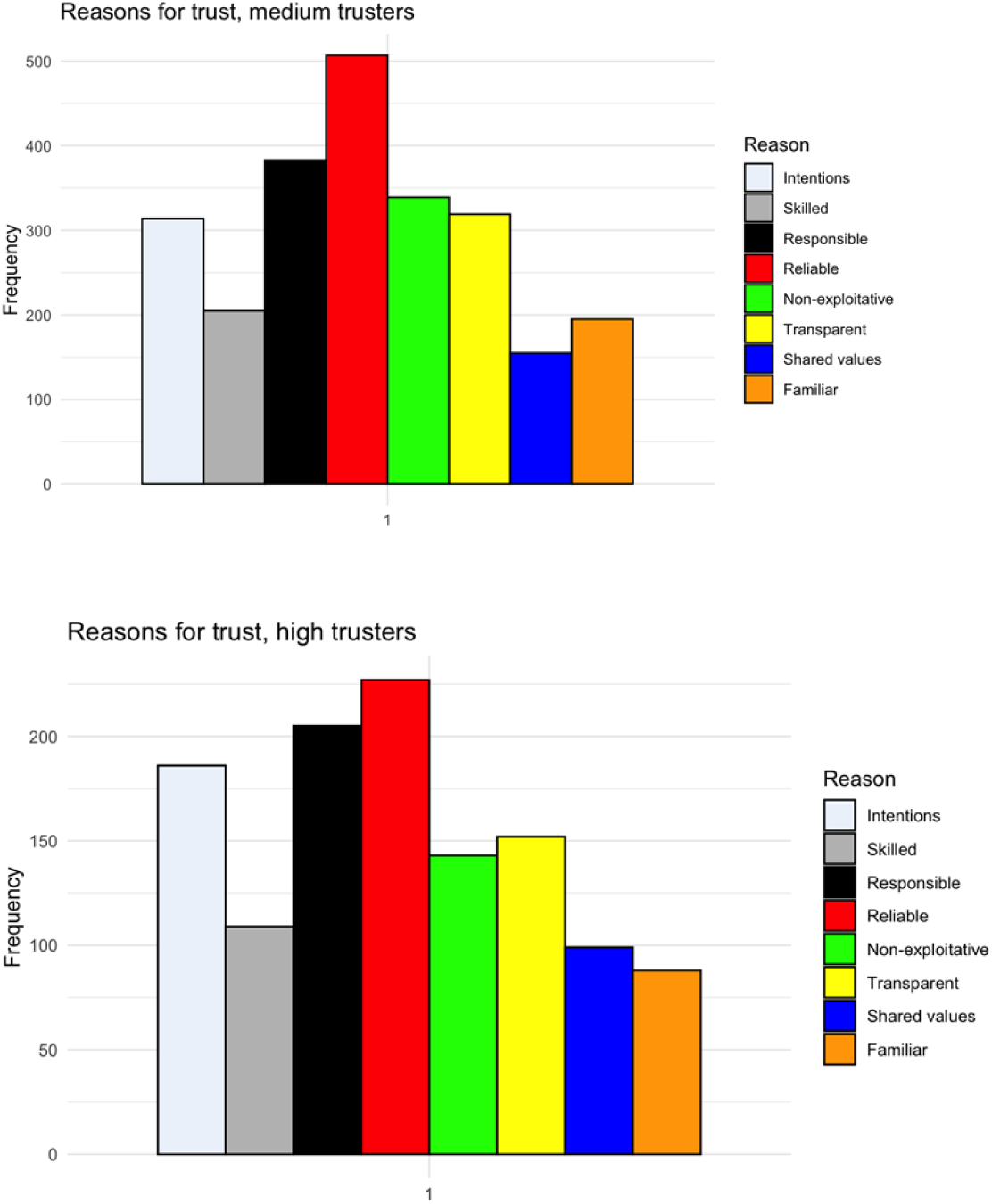
across domains, reliability was a highly rated reason for the placement of trust, though low trustors placed the most emphasis on responsibility. Shared values and familiarity, by contrast, were consistently chosen as the least important reasons across the three groups.

Figures 8, 9, and 10 give the relationships, respectively, between composite trust and previous use of health technology tools, trust in tech companies and the likelihood of using a health technology tool, and trust in the NHS and the likelihood of using a health technology tool. The first of these relationships was strong and negative; the second two were strong and positive. We discuss the compatibility of these findings below.

**Figure 8:**
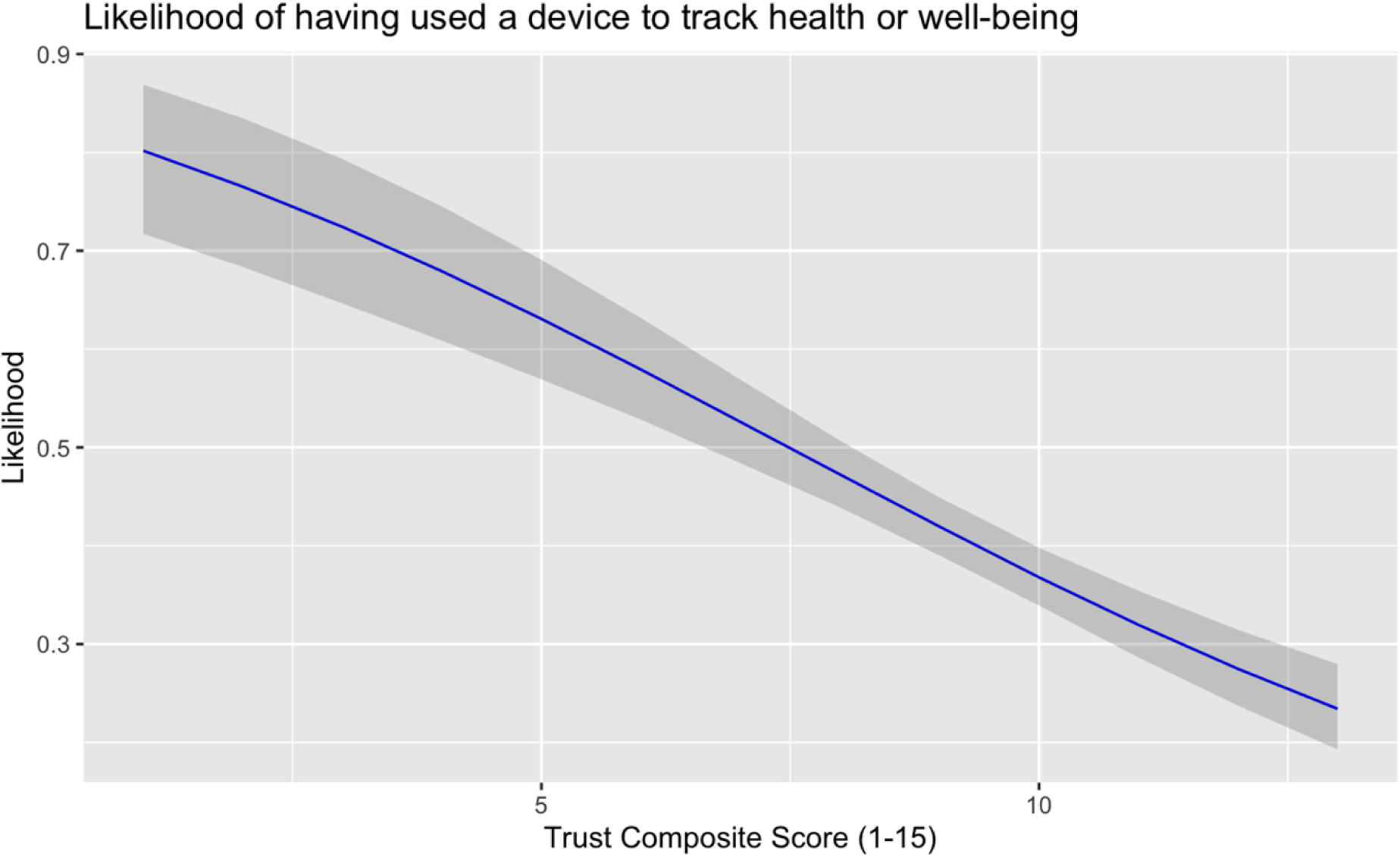
A higher composite trust score inversely correlated with the likelihood of having previously used a device to track health or well-being.

**Figure 9:**
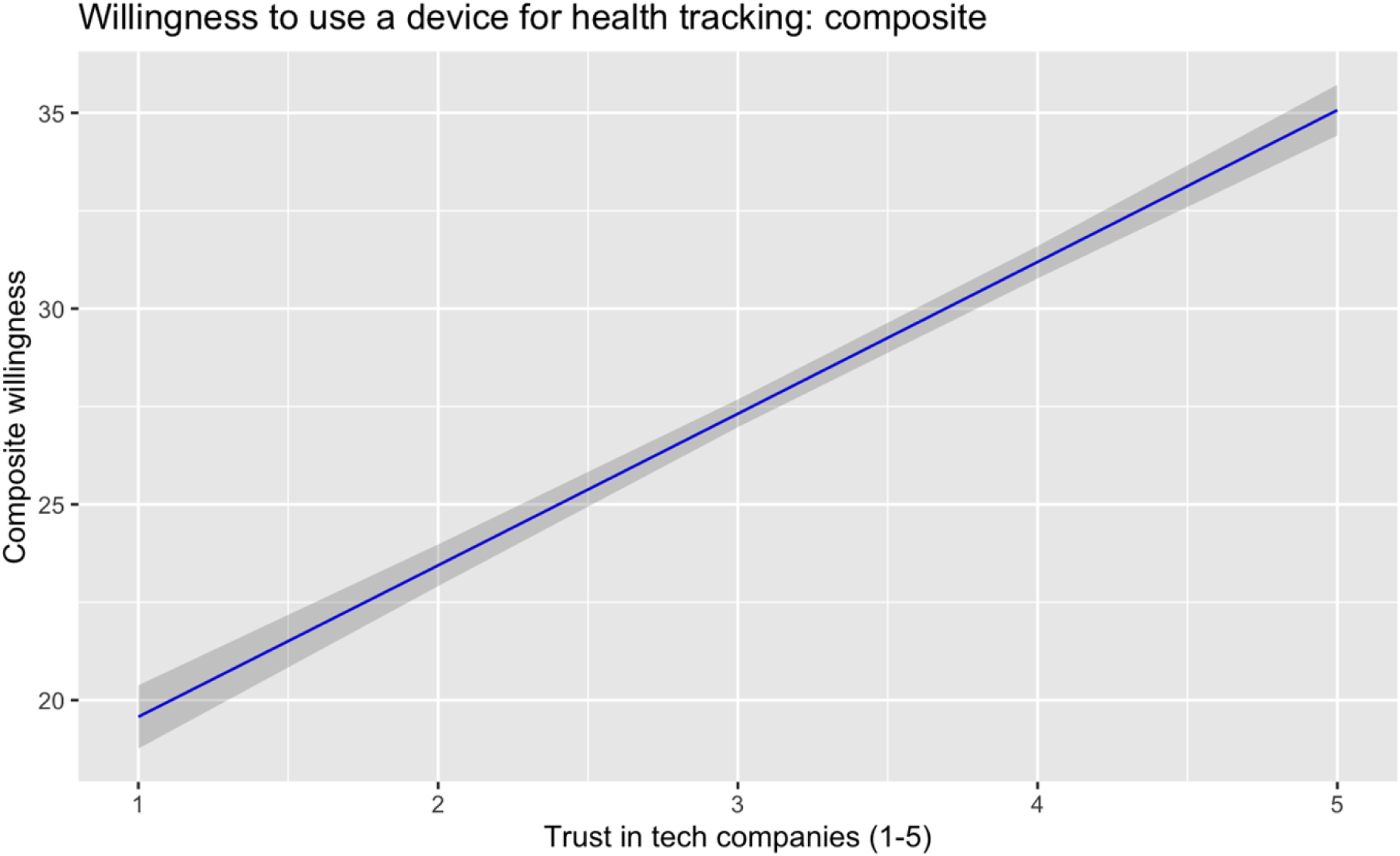
A higher trust in tech companies predicted a greater willingness to use a device for health tracking.

**Figure 10:**
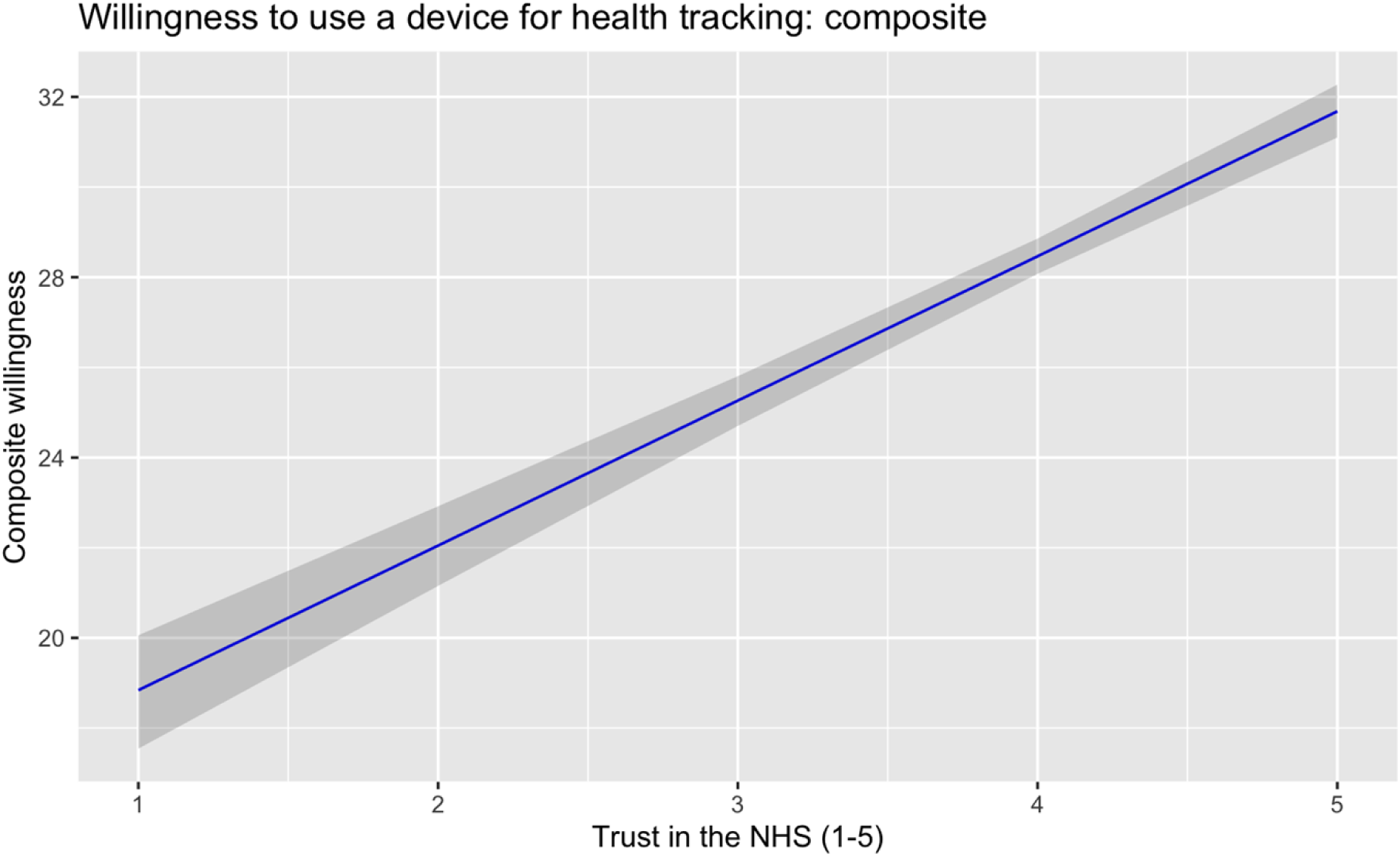
A higher trust in the NHS predicted a greater willingness to use a device for health tracking.

## Discussion

In considering a range of different reasons for trust, we have endeavoured to translate between conceptual accounts of trust and trustworthiness and empirical work. However, our central findings suggest that reasons for the placement of trust in practice are fluid and grounded in the trustor’s relationship to the trustee. For example, while several reasons for trust, such as perceived reliability, may be rated by members of the public as important with regard to health data, in specific domains, such as whether family should be trusted with one’s health data, familiarity may be the primary reason for trust placement.

The situated and contextual nature of trust relationships suggested by this finding accords with qualitative and ethnographic work that shows that trust emerges contextually by domain, and depending on the relationship between trustor and trustee (cf Cohn, 2015; Atutornu et al., 2022). This presents a challenge to efforts to establish accounts of trust that are not only normative but also functionally relevant. The question of how best to place trust, then, is more complex than assuming individuals will trust others who demonstrate trustworthiness generally: the manner in which trustworthiness must be demonstrated is a function of the trust relationship and the broader information environment.

Within a domain such as health data sharing, for example, about which there is a large published literature and body of media reports (see Baines et al., 2024b for a review), how individuals decide whether a given institution is trustworthy will depend not only on factors like the institution’s overall reputation, but the qualities that a specific trustor believes this type of trustee should have.

Accordingly, while members of the public may have specific concerns about the potential problems that could emerge from organisations having access to health data (Lounsbury et al., 2019), the placement of trust may be driven not entirely by these concerns, but rather the trustor’s qualities in relation to the trustee. Trustworthiness with health data, in this regard, is an underlying quality (Goodman & Milne, 2024) that manifests and is perceived differently under different conditions: trusting my family may be a question of familiarity, trusting the NHS may be a question of perceived responsibility, and trusting corporations may be a question of perceived transparency.

When participants are grouped into clusters of low, medium, and high trust in relation to the three types of trustee, there are, however, consistent qualities that are rated highly or lowly as reasons for placing trust (see also Milne et al., 2019b). Responsibility and reliability thus persist as primary reasons across groups, while shared values and familiarity are less important — despite familiarity being the primary reason chosen for trust in family members. Evaluating the reasons for trust absent of understanding the relationship between trustor and trustee may, therefore, obfuscate important elements of why trust is placed.

Interestingly, the consequences of trust suggested by these findings are complicated. Thus, whether an individual self-rates as trusting across the family, NHS, and health technology company domains may be predictive of intended behaviours, such as previous use of health technology tools or apps, but not of existing behaviours. One possibility is that this reflects the different social and political positions associated with digital health. Over the last 15 years, the widespread adoption of digital health tools has been closely associated with the move towards self-tracking and the ‘Quantified Self’ movement. This associates digital health devices with individual empowerment, responsibility, liberation from corporate or system control (Sharon & Zandbergen, 2017), and ‘soft resistance’ to data aggregation (Nafus & Sherman, 2014). Adoption of health technologies may therefore be perceived to be in tension with institutionalised and solidaristic forms of healthcare (McFall, 2019). In some cases, for example, the extensive adoption of digital self-monitoring is celebrated for its ability to permit forms of self-care that cannot be provided by health systems, or that enable individual control of data about health (Sharon and Zandbergen 2017). As a result, we might expect those individuals who conform most closely to the model of those who “generate data about themselves for themselves” (Catlaw & and Sandberg, 2018) to lie at an extreme of high adoption and low trust in institutions involved in healthcare. Conversely, those who have most trust in the wider health system may feel a greater willingness to rely on it, and lower need to monitor their own health.

In contrast, the willingness of ‘high trustors’ to use such tools in the future if proposed by the NHS, reflects a differing logic of adoption. In this case, digital tools are part of, rather than in addition or opposition to, health systems. This disjuncture between existing and future adoption poses interesting questions and challenges for solidarity-based efforts to ground the introduction of digital and data-driven technologies into health systems (Horn & Kerasidou, 2020).

It is important to note, however, that there are other barriers to the use of these tools that do not involve trust. These barriers may include the potential benefits these tools might offer, or the lack thereof. If, for example, an individual is willing to entrust personal health data to a company, but does not believe the benefits of using the tool are sufficient to purchase one, trust is irrelevant to use.

The findings suggest a need for a conceptual account of trust that accounts for domain-specific relationships such as those in the context of health data, and a series of questions that are pertinent to determining whether trust is likely to exist between two parties. These may include:

– What is the trustor’s relationship to the potential trustee?
– Is the trustor in a position to exploit the trustee without severe repercussions?
– What benefits may the trustor gain from sharing information with the trustee?

Answering these questions in a given relationship will help to determine whether trust is or ought to be placed, and further highlight, following the results here, the reasons likely to be important to the trustor in making a decision about the trustee.

We suggest, overall, that our results illustrate the value of working towards accounts of trust that captures its contextual and relational nature. This would enable a better understanding of how trust is placed in practice in the context of health data and more broadly. More data, furthermore, are necessary to uncover the varying reasons people have for placing trust in varying kinds of trustees, accounting for complex interplays, such as whether trustees are individuals or institutions.

## Data Availability

All files and code related to this study are available at https://github.com/jonathanrgoodman/SPACE.

## Works cited

1. Atutornu, J., Milne, R., Costa, A., Patch, C., & Middleton, A. (2022). Towards equitable and trustworthy genomics research. EBioMedicine, 76, 103879. 10.1016/j.ebiom.2022.103879

2. Baier, A. (1986). Trust and Antitrust. Ethics, 96(2), 231–260.

3. Baines, R., Stevens, S., Austin, D., Anil, K., Bradwell, H., Cooper, L., Maramba, I. D., Chatterjee, A., & Leigh, S. (2024a). Patient and Public Willingness to Share Personal Health Data for Third-Party or Secondary Uses: Systematic Review. Journal of Medical Internet Research, 26(1), e50421. 10.2196/50421

4. Baines, R., Stevens, S., Austin, D., Anil, K., Bradwell, H., Cooper, L., Maramba, I. D., Chatterjee, A., & Leigh, S. (2024b). Patient and Public Willingness to Share Personal Health Data for Third-Party or Secondary Uses: Systematic Review. Journal of Medical Internet Research, 26, e50421. 10.2196/50421

5. Bürkner, P.-C., Gabry, J., Weber, S., Johnson, A., Modrak, M., Badr, H. S., Weber, F., Ben-Shachar, M. S., Rabel, H., & Mills, S. C. (2022). brms: Bayesian Regression Models using ‘Stan’ (Version 2.18.0) [Computer software]. https://CRAN.R-project.org/package=brms

6. Catlaw, T. J., & and Sandberg, B. (2018). The Quantified Self and the Evolution of Neoliberal Self-Government: An Exploratory Qualitative Study. Administrative Theory & Praxis, 40(1), 3–22. 10.1080/10841806.2017.1420743

7. Cohn, S. (2015). ‘Trust my doctor, trust my pancreas’: Trust as an emergent quality of social practice. *Philosophy*, Ethics, and Humanities in Medicine, 10(1), 9. 10.1186/s13010-015-0029-6

8. D’Cruz, J. (2019). Humble trust. Philosophical Studies: An International Journal for Philosophy in the Analytic Tradition, 176(4), 933–953.

9. Felzmann, H., Villaronga, E. F., Lutz, C., & Tamò-Larrieux, A. (2019). Transparency you can trust: Transparency requirements for artificial intelligence between legal norms and contextual concerns. Big Data & Society, 6(1), 2053951719860542. 10.1177/2053951719860542

10. Ghafur, S., Dael, J. V., Leis, M., Darzi, A., & Sheikh, A. (2020). Public perceptions on data sharing: Key insights from the UK and the USA. The Lancet Digital Health, 2(9), e444–e446. 10.1016/S2589-7500(20)30161-8

11. Goodman, J. R., & Milne, R. (2024). Signalling and rich trustworthiness in data-driven healthcare: An interdisciplinary approach. Data & Policy, 6, e62. 10.1017/dap.2024.74

12. Hardin, R. (2002). Trust and Trustworthiness. Russell Sage Foundation. https://www.jstor.org/stable/10.7758/9781610442718

13. Hardin, R. (2006). Trust: 10. Polity.

14. Hawley, K. (2014). Trust, Distrust and Commitment. Noûs, 48(1), 1–20.

15. Hawley, K. J. (2019). How to Be Trustworthy. Oxford University Press.

16. Holton, R. (1994). Deciding to Trust, Coming to Believe. Australasian Journal of Philosophy, 72(1), 63–76. 10.1080/00048409412345881

17. Horn, R., & Kerasidou, A. (2020). Sharing Whilst Caring: Solidarity and Public Trust in a Data-Driven Healthcare System. BMC Medical Ethics, 21(1), 1–7. 10.1186/s12910-020-00553-8

18. John, S. (2021). Science, politics and regulation: The trust-based approach to the demarcation problem. Studies in History and Philosophy of Science Part A, 90, 1–9. 10.1016/j.shpsa.2021.08.006

19. Jones, K. (2012). Trustworthiness. Ethics, 123(1), 61–85. 10.1086/667838

20. Kalkman, S., Delden, J. van, Banerjee, A., Tyl, B., Mostert, M., & Thiel, G. van. (2022). Patients’ and public views and attitudes towards the sharing of health data for research: A narrative review of the empirical evidence. Journal of Medical Ethics, 48(1), 3–13. 10.1136/medethics-2019-105651

21. Laverty, L., Jones, E., Peek, N., Oswald, M., Bozentko, K., Atwood, S., & van der Veer, S. (2024). The power and promise of transparency: Perspectives from citizens’ juries of pandemic health data sharing. Big Data & Society, 11(4), 20539517241299729. 10.1177/20539517241299729

22. Lounsbury, M. (2023). The Problem of Institutional Trust. Organization Studies, 44(2), 308–310. 10.1177/01708406221131415

23. Lounsbury, O., Roberts, L., Goodman, J. R., Batey, P., Naar, L., Flott, K. M., Lawrence-Jones, A., Ghafur, S., Darzi, A., & Neves, A. L. (2021). Opening a ‘Can of Worms’ to Explore the Public’s Hopes and Fears About Health Care Data Sharing: Qualitative Study. Journal of Medical Internet Research, 23(2), e22744. 10.2196/22744

24. Lüdecke, D., Bartel, A., Schwemmer, C., Powell, C., Djalovski, A., & Titz, J. (2024). sjPlot: Data Visualization for Statistics in Social Science (Version 2.8.17) [Computer software]. https://cran.r-project.org/web/packages/sjPlot/index.html

25. Luhmann, N. (1979). Trust and Power. Wiley. https://www.wiley.com/en-gb/Trust+and+Power-p-9781509519453

26. Mayer, R. C., Davis, J. H., & Schoorman, F. D. (1995). An integrative model of organizational trust. The Academy of Management Review, 20(3), 709–734. 10.2307/258792

27. McFall, L. (2019). Personalizing solidarity? The role of self-tracking in health insurance pricing. Economy and Society, 48(1), 52–76. 10.1080/03085147.2019.1570707

28. Middleton, A., Costa, A., Milne, R., Patch, C., Robarts, L., Tomlin, B., Danson, M., Henriques, S., Atutornu, J., Aidid, U., Boraschi, D., Galloway, C., Yazmir, K., Pettit, S., Harcourt, T., Connolly, A., Li, A., Cala, J., Lake, S., … Parry, V. (2023). The legacy of language: What we say, and what people hear, when we talk about genomics. HGG Advances, 4(4), 100231. 10.1016/j.xhgg.2023.100231

29. Middleton, A., Milne, R., Thorogood, A., Kleiderman, E., Niemiec, E., Prainsack, B., Farley, L., Bevan, P., Steed, C., Smith, J., Vears, D., Atutornu, J., Howard, H. C., & Morley, K. I. (2019). Attitudes of publics who are unwilling to donate DNA data for research. European Journal of Medical Genetics, 62(5), 316–323. 10.1016/j.ejmg.2018.11.014

30. Milne, R., Morley, K. I., Almarri, M. A., Anwer, S., Atutornu, J., Baranova, E. E., Bevan, P., Cerezo, M., Cong, Y., Costa, A., Critchley, C., Fernow, J., Goodhand, P., Hasan, Q., Hibino, A., Houeland, G., Howard, H. C., Hussain, S. Z., Malmgren, C. I., … Middleton, A. (2021). Demonstrating trustworthiness when collecting and sharing genomic data: Public views across 22 countries. Genome Medicine, 13(1), 92. 10.1186/s13073-021-00903-0

31. Milne, R., Morley, K. I., Howard, H., Niemiec, E., Nicol, D., Critchley, C., Prainsack, B., Vears, D., Smith, J., Steed, C., Bevan, P., Atutornu, J., Farley, L., Goodhand, P., Thorogood, A., Kleiderman, E., Middleton, A., & Participant Values Work Stream of the Global Alliance for Genomics and Health. (2019a). Trust in genomic data sharing among members of the general public in the UK, USA, Canada and Australia. Human Genetics, 138(11–12), 1237–1246. 10.1007/s00439-019-02062-0

32. Milne, R., Morley, K. I., Howard, H., Niemiec, E., Nicol, D., Critchley, C., Prainsack, B., Vears, D., Smith, J., Steed, C., Bevan, P., Atutornu, J., Farley, L., Goodhand, P., Thorogood, A., Kleiderman, E., Middleton, A., & Participant Values Work Stream of the Global Alliance for Genomics and Health. (2019b). Trust in genomic data sharing among members of the general public in the UK, USA, Canada and Australia. Human Genetics, 138(11–12), 1237–1246. 10.1007/s00439-019-02062-0

33. Nafus, D., & Sherman, J. (2014). Big Data, Big Questions| This One Does Not Go Up To 11: The Quantified Self Movement as an Alternative Big Data Practice. International Journal of Communication, 8(0), Article 0.

34. OECD Guidelines on Measuring Trust. (2017, November 23). OECD. https://www.oecd.org/en/publications/oecd-guidelines-on-measuring-trust_9789264278219-en.html

35. O’Neill, O. (2018). Linking Trust to Trustworthiness. International Journal of Philosophical Studies, 26(2), 293–300. 10.1080/09672559.2018.1454637

36. Share of people agreeing with the statement ‘most people can be trusted’. (n.d.). Our World in Data. Retrieved 20 June 2025, from https://ourworldindata.org/grapher/self-reported-trust-attitudes?tab=chart&country=~GBR

37. Sharon, T., & Zandbergen, D. (2017). From data fetishism to quantifying selves: Self-tracking practices and the other values of data. New Media & Society, 19(11), 1695– 1709. 10.1177/1461444816636090

38. Sorrell, K. (2013). Pragmatism and moral progress: John Dewey’s theory of social inquiry. Philosophy & Social Criticism, 39(8), 809–824. 10.1177/0191453713494967

39. Stewart, E. (2024). Negotiating domains of trust. Philosophical Psychology, 37(1), 62–86. 10.1080/09515089.2022.2144190

40. Team, R. C. (2022). R: A language and environment for statistical computing. R Foundation for Statistical Computing. https://www.R-project.org/.

41. Wickham, H., Chang, W., Henry, L., Pedersen, T. L., Takahashi, K., Wilke, C., Woo, K., Yutani, H., Dunnington, D., & RStudio. (2021). ggplot2: Create Elegant Data Visualisations Using the Grammar of Graphics (Version 3.3.5) [Computer software]. https://CRAN.R-project.org/package=ggplot2

